# Aerosol filtering efficiency of respiratory face masks used during the COVID-19 pandemic

**DOI:** 10.1101/2020.07.16.20155119

**Authors:** Glykeria Loupa, Dimitra Karali, Spyridon Rapsomanikis

**Affiliations:** Laboratory of Atmospheric Pollution and Pollution Control Engineering of Atmospheric Pollutants, Department of Environmental Engineering, Faculty of Engineering, Democritus University of Thrace, P.O. Box 447, 67100 Xanthi, Greece

**Keywords:** Face masks, Aerosol dynamics, Particle filtration, Protective equipment, Indoor air

## Abstract

The spread of the COVID-19 pandemic, effected the imposition of personal protection measures in a large number of countries. The use of commercially available personal face masks was widely accepted as such a protective measure. Since the quality of the face masks scanned the spectrum from surgical to the home made fabric ones, it was considered appropriate to experimentally establish their effectiveness for stopping aerosol in entering the respiratory system of the bearer. Presently, only eight masks were tested with polydisperse indoor air. Their effectiveness was examined for aerosol of aerodynamic diameters of 0.006 μm to 10 μm. Of these masks, only two were effective for the whole range of aerosol. Cloth masks were found to be ineffective for the assigned task.

## INTRODUCTION

The recent pandemic of COVID-19, has been the concern of the scientific community of most disciplines. The pandemic introduced the widespread application of policies for constraining its spread (Fineberg, 2020; National Academies of Sciences and Medicine, 2020; World Health Organization, WHO, 2020). Apart from lockdowns of cities, public services and whole countries, individual measures suggested and enacted by state decrees, included the use of personal respiratory face masks (MacIntyre *et al.*, 2015; Centers for Disease Control Prevention, CDC, 2020).

The new virus SARS-CoV-2, as all the viruses in the atmosphere are particles floating in the air (aerosol). Aerosol (or airborne particulate matter, PM) is defined by the International Union of Pure and Applied Chemistry, IUPAC as: “*Mixtures of small particles (solid, liquid, or a mixed variety) and the carrier gas (usually air); owing to their size, these particles (usually less than 100* μ*m and greater than 0.01* μ*m in diameter) have a comparatively small settling velocity and hence exhibit some degree of stability in the earth’s gravitational field”* (Calvert, 1990). Viruses are a category of aerosol of biological origin and referred to as bioaerosol (Douwes *et al.*, 2008; Yao, 2018). Viruses, such as SARS, H1N1 and MERS-CoV, pose distinct health risks, in addition to the known aerosol health effects (Yao, 2018). Their transmission from person to person through the atmosphere is a pathway of infection that is not well explored, although a very important one, especially for the indoor environments (Nazaroff, 2016; Yao, 2018; Anderson *et al.*, 2020; Fears *et al.*, 2020).

Bioaerosol has similar dynamics with all the airborne particles and the prevailing parameter that characterises its fate in the atmosphere is its size. How far these particles can be transmitted (emitted from coughing, or sneezing or even from talking from infected persons) and for how long they remain suspended, depends on their size (Lee *et al.*, 2019). The gravitation settling is more significant for the larger particles, prohibiting them to be transmitted to long distances. However, exhaled droplets become dry particles in the air, hence their size is reduced (fast) (Morawska *et al.*, 2009; Hinds, 2011). For the smaller particles, turbulence is the dominant dispersion mechanism, enabling them to travel long distances from their source and for a long time.

Hence, the health effects and the possible protective measures against aerosol containing viruses, depend on several critical parameters, such as the particle size, the aerosol atmospheric number concentration in different sizes, the aerosol atmospheric lifetime and the atmospheric lifetime of the virus associate with it (Guzman, 2020; Mutuku *et al.*, 2020). Such parameters will determine how far viruses can be transmitted depending on the range of the sizes of the airborne particles carrying the viruses, how deep in the respiratory system can penetrate if inhaled and in the end their infectivity (Kunkel *et al.*, 2017).

Morawska *et al.*, (2009) reported that the majority of particles emitted during all expiratory activities of humans have diameters below 0.8 μm, and their size distribution had one or more modes. A recent publication for two Wuhan hospitals (Liu *et al.*, 2020), indicates that the most abundant, in number, viral particles have aerodynamic diameter between 250 nm and 500 nm.

Face masks have been used for years to protect the public from aerosol or bioaerosol and the filtration efficiency of individual masks has been examined in several studies (for example (Rengasamy *et al.*, 2010; Mueller *et al.*, 2018). This efficiency depends on the material of the mask, its design (e.g. with exhaust valve or not), the face velocity of the aerosol that impacts on the mask and the fitting of the mask to the person’s face (Rengasamy *et al.*, 2010; Steinle *et al.*, 2018; Dbouk and Drikakis, 2020; Konda *et al.*, 2020). Finally, the aerosol filtering efficiency of a face mask protecting from viruses, has to be examined in relation to the spectrum of the sizes of particles caring viruses (internally or externally mixed) that travel in the air and reach our respiratory system (Drossinos and Stilianakis, 2020; Konda *et al.*, 2020).

Due to high demand for masks worn by the public and the shortage in their supply during the COVID-19 pandemic masks of unknown efficiency and quality appeared in the market.

In the present work, the filtration efficiency of eight commercially available face masks was examined under near realistic conditions as a function of the different sizes of aerosol found in an indoor environment which ranged between 6 nm and 10 μm. Some theoretical considerations on the airborne transmission of the exhaled respiratory droplets and the examination of the filtration efficiency of the 8 masks in relation with aerosol size distribution is also determined.

## METHODS

Eight (8) respiratory face masks for use by the general public were tested for their aerosol filtering efficiency in an indoor environment of the campus of the Democritus University of Thrace, Xanthi, Greece, during May 2020. These masks were commercially available worldwide and at a price that most can pay, i.e. between 0.8 and 5 Euro. In Greece, they are sold in pharmacies, but one can also purchase them from the internet. Five of them are 3-layer masks (referred to as M1, M2, M3, M4, M5), one is a KN95 mask with “one way valve” (named as M6). Two masks referred to as F1 and F2, are fabric masks. The characteristics of the tested masks are presented in Table1, as they are provided on the label of their packaging. Note that only half of them have a brand name and their photos are depicted in the graphical abstract of the present work.

The experiments were conducted inside an indoor environment which was naturally ventilated with an air exchange rate of 0.8-1.1 h^-1^ for a volume of 150 m^3^. The instrumentation used comprised of two supplementary, in series, set ups. A light scattering particle counter, PROMO 2000 and an SMPS –CPC particle counter were used (Palas®, Karlsruhe, Germany). For details see the Supplementary Material.

Each mask was fitted directly on the inlets of the instruments (see Fig. 1). Data were collected for 30 min with the inlets open in the air of the room (background) and for the next 30 min with the inlets covered with a mask. For each of the eight tested type of mask, six replicates were conducted. Occasionally, the inlet was moved to a different area of the mask, to ensure homogeneous coverage of the penetration area. The instrumental rate of sampling air at 0.5 l min^-1^ for the SMPS-CPC and an inlet tubing 3/8” or ca 6 mm diameter, resulted in a face velocity of the entering aerosol (PM_2.5_) of 0.3 m s^-1^. The face velocity of the air entering the mask of an average person that has a breathing rate of ca 8 l min^-1^ and an average area for two nostrils of ca 700 mm^2^ is calculated to be ca 0.38 m s^-1^ per nostril or 0.19 m s^-1^ per breath (Schriever *et al.*, 2013). The later values are similar to the instrumental sampling “face velocity”. The sampling face velocity directly affects the separation factor N_s_, which in turn is directly related to a filter efficiency: N_s_= ρD2V/18μD_b_; where ρ= particle density, D = particle diameter D_b_ = fiber diameter and μ = kinematic viscosity of air (Langmuir and Blodgett, 1946; De Nevers, 2010). If the other parameters of the above equation remain constant, then the critical parameter in the evaluation of filter efficiency in near realistic conditions, is the face velocity of the air stream that carries the bioaerosol and “hits” the filtering area.

**Fig. 1.**
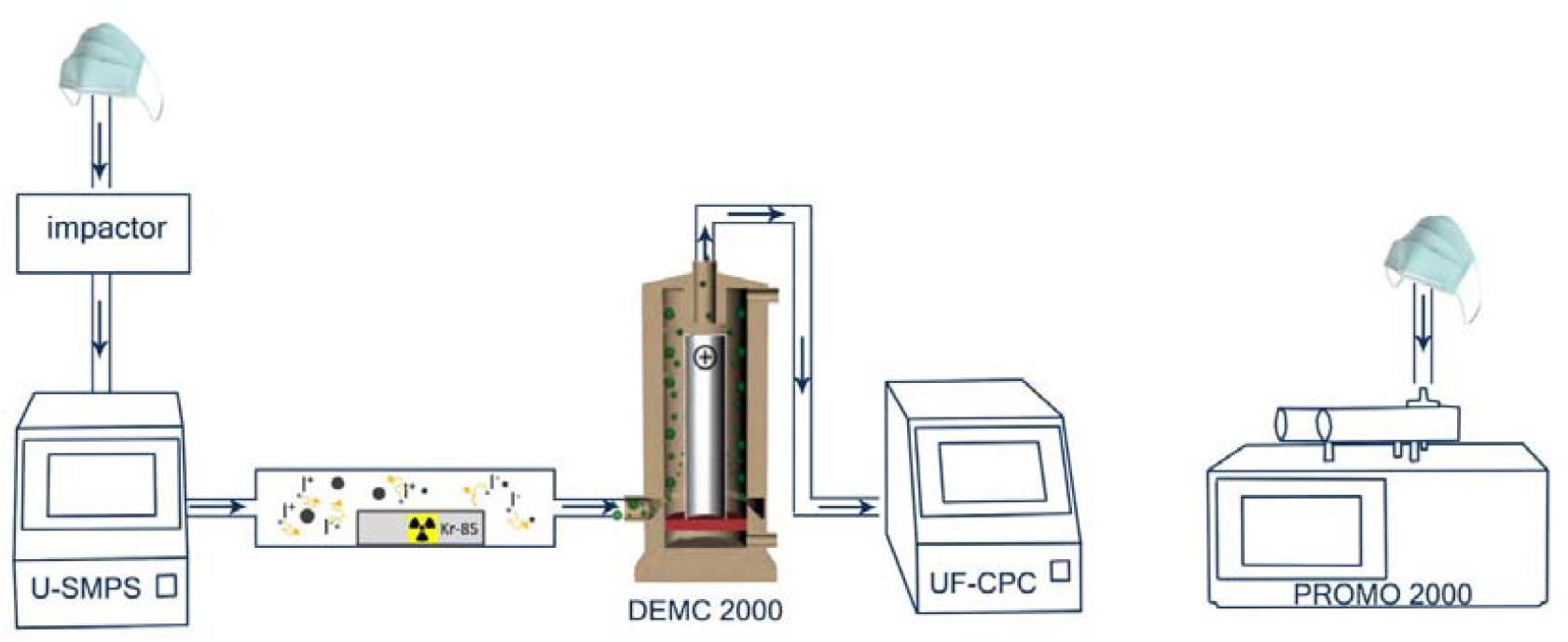
The aerosol sampling instrumentation where U-SMPS denotes the Universal Scanning Mobility Particle Sizer control unit; DEMC denotes the Differential Electrical Mobility Classifier; UF-CPC denotes the Universal Fluid - Condensation Particle Counter (butanol in the present case) and PROMO 2000 denotes the Optical Scattering Particle Counter.

The eight (8) commercially available masks were examined for their efficiency in withholding airborne particles of sizes 0.006 to 10 μm in 102 size bins. This filtration efficiency (FE) was calculated for the number concentration of particles, by the following equation:

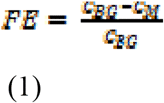

where, in each size bin, the C_BG_ (BG= background) was the average particle number concentration before putting the mask on the inlet, whilst C_M_ is the average particle concentrations per size bin when the instrument sampled indoor air that passed through the mask (M= filtered through the mask).

## RESULTS AND DISCUSSION

### Filtration efficiency of the different masks

An average PM size distribution before putting the mask on the inlet of the instrument and when the mask was in place are presented in Fig. 2, for two examined masks, as an example of low and high FE. The particle size distributions in the room indicate that the larger number concentrations of the sampled aerosol can be found in the 20-350 nanometre scale, an area where peak concentration of SARS-CoV-2 particles were observed (Liu *et al.*, 2020).

**Fig. 2.**
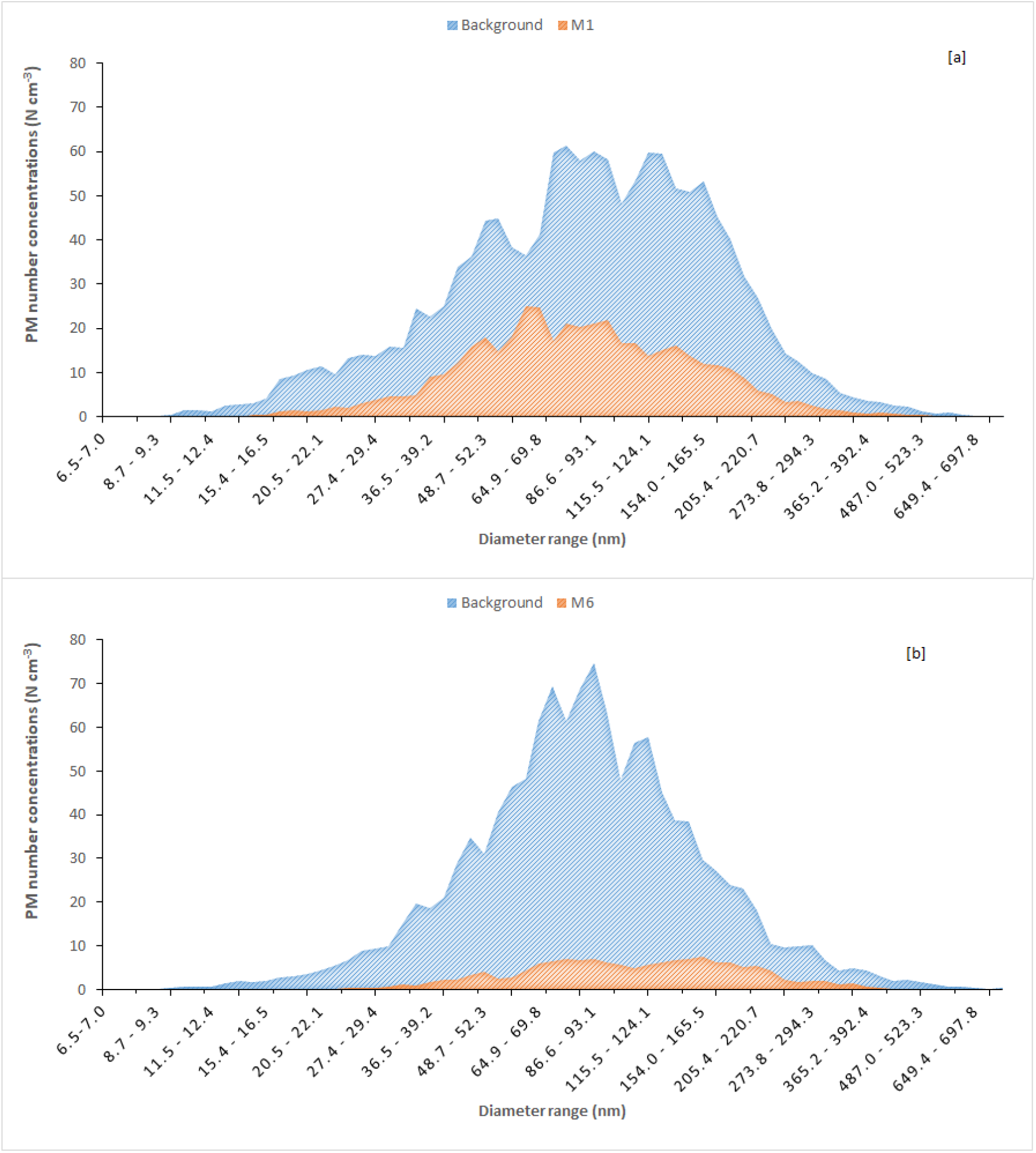
Average PM number concentrations for a period of 30-min as recorded before and after filtration through the mask. Diagram (a) indicates low efficiency and (b) high efficiency across the size bin range indicated.

Fig. 3 compares the FE for all the masks under study. The 102 size bins were reduced to 14 bins for each examined mask for reasons of clarity. In the Fig.3, the superimposed rectangle shadowed part, aims to highlight the size range where the higher number concentration of SARS –CoV-2 can be found (Liu *et al.*, 2020). The simple cloth mask F1 (with two layers of cotton, like the home made masks) has the lowest FE from all, in agreement with other studies (Rengasamy *et al.*, 2010; Davies *et al.*, 2013; Cherrie *et al.*, 2018; Konda *et al.*, 2020). The addition of activated carbon in a cotton mask, the F2 in our case, improves significantly its FE (Fig. 3(a)). The FE of the masks M1, M2 (no brand name) is between F1 and F2, but they exhibited a larger FE than F1 and F2 for particle sizes above 800 nm. In the Fig.3(b) the masks with better FEs than the masks in Fig 3a. are presented. The M6 has FE above 90% in all size bins. The M6 is a KN95 mask similar with the standard N95 mask in US. It appears that apart from the M6, the other masks are more or less permeable to the aerosol in the range 250-500 nm. Note that that the masks that public wears, very rarely fit well in their face. This leads to a further significant reduction of the mask effectiveness and hence to the protection against viruses (Cherrie *et al.*, 2018; Dbouk and Drikakis, 2020; Konda *et al.*, 2020; Peric and Peric, 2020).

**Fig. 3.**
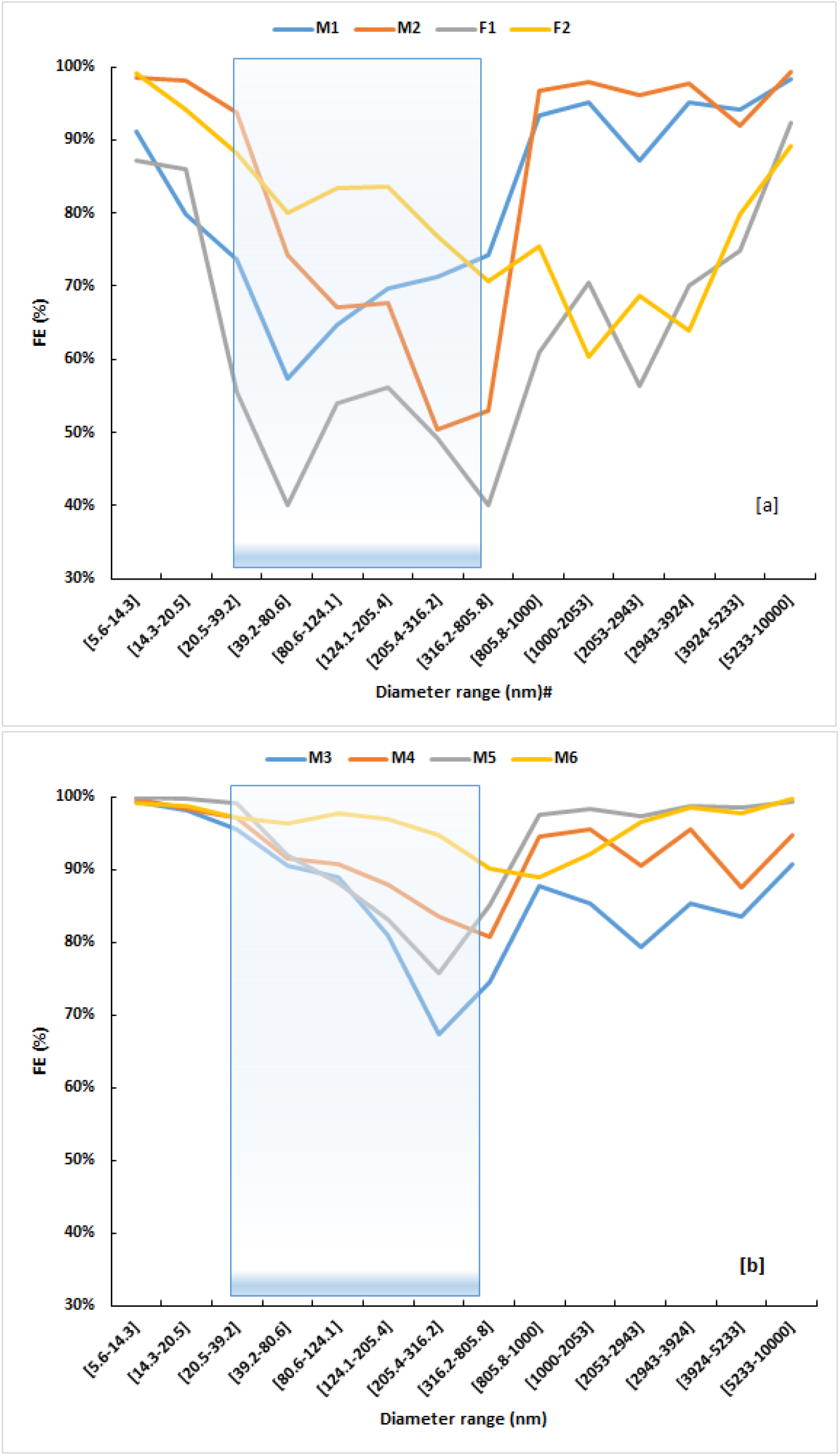
Filtration Efficiency for all the masks under study for 14 size bins. The mask nomenclature is the same as noted in Table 1.

The present findings are consistent with the reported results in other studies that have examined the FE in relation with particle size distribution, for example the study of Konda *et al.*, (2020) or older studies such as the study of (Mueller *et al.*, 2018), despite the differences in the setup of the experiments of each study, for example artificial aerosol versus real indoor aerosol.

**Table 1.** Summary of the characteristics of each tested mask.

| Mask | Brand | Description |
| --- | --- | --- |
| **M1** | No available name | Face mask with stretchable elastic ear loops<br>Soft-breathable-skin friendly, Effective protection<br>Comfortable material<br>BFE 95% |
| **M2** | No available name | Hygiene & Protection<br>Face masks 3-Ply Disposable, PP non-woven |
| **M3** | HYGOSTAR | Face mask with ear loops, green, 3-ply, nickel-free<br>Materials- outer layer: PP (polypropylene non-woven) fabric-like<br>- middle layer / filter: melt blown<br>- outer layer: PP (polypropylene non-woven)<br>excellent filter performance > 99% BFE<br>non sterile, lint-free, skin-friendly<br>integrated nose bridge, adjustable anatomically |
| **M4** | YI HU KANG<br>HEALTHY | Medical Face Mask Ear Loop<br>3 layer protective filtering ~ Anti-fog, Anti-droplet, Dust and Effective<br>Filters bacterial<br>• Inner Layer: Skin-Friendly and comfortable<br>• Filter Layer: Filter oily and non-oily particles, antibacterial material<br>• Outer Layer: Block larger particles<br>• BFE=>95%<br>Material composition: Inner and outer premium polypropylene non-woven fabric, Middle antibacterial filter layer, 3-layer thickened protection design |
| **M5** | ASEPTA | Surgical Face Mask Ear Loop<br>For single use<br>Hypoallergenic - 3ply<br>Fiberglass free<br>>95% B.F.E.<br>Long integrated nose-piece<br>Antibacterial filter between layers |
| **M6** | HENGHAO | Disposable 3 Layer Medical Face Mask with elastic ear loops<br>KN95<br>(*Chinese standards for masks nearly equivalent on the features with the N95 based on US standards*)<br>BFE>95%<br>Effectively resists PM<sub>2.5</sub> and protects against dust, smoke and microorganisms |
| **F1** | Greek product<br>No available name | Face mask with stretchable elastic ear loops<br>100% cotton |
| **F2** | Greek product<br>No available name | Face mask with stretchable elastic ear loops<br>Cotton mask with activated carbon<br>Multiuse |

### Aerosol dynamics

There exists a number of recent publications that discuss and confirm the transmission of viruses and of SARS-CoV-2 via the atmosphere (Yu *et al.*, 2004; Kim *et al.*, 2016; Kutter *et al.*, 2018; Tellier *et al.*, 2019; Anderson *et al.*, 2020; Asadi *et al.*, 2020; Fears *et al.*, 2020; Lednicky *et al.*, 2020; Prather *et al.*, 2020). The major parameters that were theoretically discussed below concerning this transfer process, are the exit stopping distance of aerosol from the respiratory processes, the gravitational settling velocity of aerosol (GSV) and the turbulence of the environment that this transfer/infection may take place. The GSV of particles in free air that are relevant to the human respiratory system, after incorporating the Cunningham Correction (slip) Factor can be in simple terms be calculated via the following equation, extracted from the tables of the following publications (Crowder *et al.*, 2002; Kulkarni *et al.*, 2011), (see also Supplementary Material).

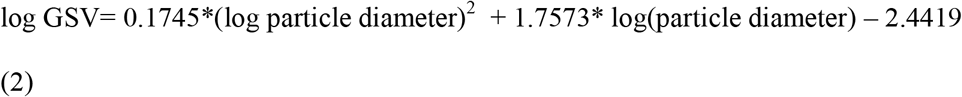

Aerosol exhaled, from the respiration process, breathing, talking, sneezing or coughing, acquires a horizontal travel distance that depends on its diameter, density and exit velocity as it appears in Equ. (3) (de Nevers 1995; Hinds, 201).

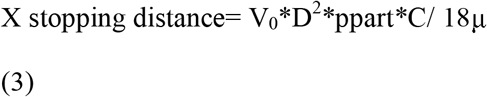

where X is the Stokes law stopping distance, Vo is the exit velocity of the particle, D is its aerodynamic diameter, ppart is its density, C is the Cunningham Correction Factor (unity for large particles) and μ is the kinematic viscosity of air.

Literature reports indicate that sneezing and coughing velocities of any density aerosol vary between 5 and 30 m s^-1^ (Stelzer□ Braid *et al.*, 2009; Mittal *et al.*, 2020). Hence, the Stokes law stopping distance of large aerosol can be calculated. If for example these aerosol have a diameter of 50 μm and a density of 2 kg m^-3^, the result is 46.3 cm, ignoring gravitational forces. If sneezing and coughing aerosol is comprised of spherical aerosol and to a lesser extent by coagulated filaments, it can travel for ca. 0.5 m. Practice, from time lapse and fast frame recording cameras, indicates a slightly larger plume (Hsiao *et al.*, 2020).

Another factor that enters the equation of exhaled human plume is the behaviour of the aerosol in an environment with relative humidity lower than that of the human lungs. The time it takes for a single particle to dry depends on its diameter and density at certain temperature and humidity as shown in Equ. (4) (Hinds, 2011).

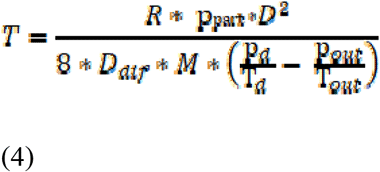

where R= the universal gas constant; p_part_=2000 kg m^-3^; diameter of particle D= 5×10^−5^ m; diffusion coefficient for water D_dif_ = 2.4×10^−5^ m^2^ s^-1^; M= molecular mass; temperature outside the particle T_out_=293 K; pressure outside the particle p_out_= 1170 Pa; T_d_ on the surface of the particle depends on the T_out_ and the saturation ratio of the particle at 293 K and finally p_d_ is the vapour pressure of water at 293 K. Equ. (4) is valid without correction for particles larger than 2 μm. Hence, an aqueous particle of 50 μm diameter after exhalation to 50% ambient relative humidity has an evaporation time of ca. 12 seconds.

Theoretical considerations may also allow the calculation of the coagulation time of aerosol to lower number concentrations of larger coagulated particles. However, these calculations are not trivial and with large uncertainties in the outcome. It appears that only measurements in near real environmental conditions will assess the atmospheric lifetime of the SARS-CoV-2 virus in a specific examined environment. It may, thence, be possible to categorise its lifetime in “generalised” environmental conditions. The above considerations indicate that the size distribution of SARS-CoV-2 virus carrying aerosol, may spread across the size range important for the human health. Hence, masks that available to the public must be efficient across the whole of this size range 10 nm to 25 and 50 μm.

Another, unanswered and difficult to answer question remains the “infection dose” to the recipients. Atmospheric concentrations of externally or internally mixed size segregated active aerosol must be related to the time of exposure, a parameter directly related to aerosol lifetime.Aerosol science has an important role to play in the understanding of the spread of viruses and the selection of protective measures.

## CONCLUSIONS

There exists ample evidence that viruses are transmitted via the atmosphere and this may be the main transmission pathway. Deposition of exhaled viral aerosol on surfaces and transmission after contact with surfaces, is a secondary but also important pathway. In the present work we examined the efficiency of face masks, available to the public at low cost, for stopping aerosol in the size range of 0.006 μm to 10 μm, in entering the individual’s respiratory system. From the aerosol science point of view, real aerosol was tested in our experiments, representative of a naturally ventilated typical indoor environment. Only surgical masks of known origin, displayed an acceptable efficiency across the aerosol range. Specifically, since the size of the airborne SARS-CoV-2 virus displays a large number concentration around 100-350 nm before its atmospheric aging, i.e. accumulation and coagulation, it was established that masks should be highly efficient at this aerosol nanoscale range. Furthermore, the instrumental sampling face velocity for the masks tested was similar to the “face velocity” of air entering the mask at a typical human breathing rate. This ensured that our experiments were not biased or erroneous because of “different/varying” face velocity parameter that affects the separation number (Ns) of aerosol for a target (mask material) efficiency. The M6, a KN95 mask was the most efficient across the examined range. Market available cloth masks were inefficient at any aerosol size range. Epidemiological studies, the resulting damage due to the pandemic is established leaving a number of unknown parameters of the “exactly how”, unanswered. The application of aerosol science and technology in infection control with extended, dedicated and organised research will provide defensive measures against the next wave of virus infection. One such very important measure (policy) would be the use by the public of surgical masks of known, tested and standardized quality.

## Data Availability

no available data

## ACKNOWLEDGMENTS

The present work was funded by Democritus University of Thrace (Greece) funds.

## DISCLAIMER

Reference to any companies or specific commercial products does not constitute their endorsement.

